# Quantitative fluorescence in situ hybridization (FISH) of magnetically confined bacteria enables rapid determination of early-stage human bacteremia

**DOI:** 10.1101/2021.02.19.21251962

**Authors:** Min Seok Lee, Hwi Hyun, Inwon Park, Sungho Kim, Dong-Hyun Jang, Seonghye Kim, Jae-Kyeong Im, Hajin Kim, Jae Hyuk Lee, Taejoon Kwon, Joo H. Kang

## Abstract

The current diagnosis of bacteremia mainly uses blood culture, which is insufficient to offer rapid and quantitative determination of pathogens in blood. Here, we report a quantitative and sequential multiplexed fluorescence in situ hybridization in a microfluidic device (µFISH) that enables early and rapid (2-hour) diagnosis of bacteremia without prior blood culture. Mannose-binding lectin-coated magnetic nanoparticles enrich a broad range of pathogens, and µFISH enables identification and quantification of the magnetically confined bacteria. We detect *Escherichia coli* (*E. coli*) and measure their relative proportions to universal bacteria levels in the bacteremic blood of a porcine model and human whole blood collected from *E. coli*-infected patients, which was elusive with the conventional bacteremia diagnosis methods. Thus, µFISH can be used as a versatile tool to rapidly identify pathogens and further assess the number of both culturable and non-culturable bacteria in biological and environmental samples.

## 1. Introduction

The presence of bacteria in the bloodstream often provides decisive evidence of systemic infection in patients. Thus, the identification of pathogenic bacteria, as well as the determination of their presence in blood, is an essential step toward the successful treatment of individuals infected with bacteria. The current gold standard method for diagnosing bacteremia mainly relies on blood culture, which is incapable of providing immediate results to clinicians ^[1]^ and often leads to false-negative outcomes ^[2]^. The rapid determination and identification of bacteria in blood are important because the delayed administration of antibiotics results in increased mortality ^[3,4]^, and prompt care for infection is frequently required for patients in the emergency department and the intensive care unit (ICU) of hospitals. In addition, given that the pathogen load is proportional to the mortality of the patients ^[5]^, it is critical to gain quantitative information of bacteria in the blood and collect taxonomical evidence of bacteria largely obtained by blood culture. While time-to-positivity (TTP) of blood culture has been considered to be a prognostic tool for a limited range of bacterial species, such as *S. aureus, E. coli*, and *S. pneumonia* ^[6–10]^, it has not been applied widely to a range of bacteria because correlations between TTP data of certain bacterial species and mortality of the patients turned out to obscure in several clinical studies ^[11]^. Despite these drawbacks of the blood culture method, it is still the most widely used clinical laboratory methods to determine causative bacteria in the blood due to the lack of complementary approaches.

Identification of bacteria following expansion in blood culture is another crucial procedure for treating the bacteria-infected patients properly. Mass spectrometry (MS) ^[12,13]^ and polymerase chain reaction (PCR) ^[14,15]^ have been mainly incorporated in the diagnostic procedure to identify bacteria in the blood. However, they are often available only after significantly expanding bacteria in biological samples *in vitro*, which is time-consuming and unavailable for certain bacterial species due to insufficient growth rates in a standard culture condition. Although the commercially available MS and PCR-based diagnostic systems allowed to work directly with whole blood, eliminating the most time-consuming step for the bacterial culture ^[16,17]^, their clinical impact is still contentious due to the variable sensitivity and specificity arising from the presence of several inhibitors (human DNA, iron, and heparin) and complex background signals in blood ^[18]^.

Fluorescence in situ hybridization (FISH) has also permitted the unique capability in a cytogenetic analysis by using fluorescent probes that are hybridized with complementary nucleic acid sequences, which brought considerable impact on cell biology and genomics ^[19– 22]^. Its quantitative analytical ability in measuring fluorescence signals in a cell (Q-FISH) also enabled quantitation of gene expression and genomic elements in cells or tissues ^[23–25]^. Later, a modification of FISH combining with flow cytometry (Flow-FISH) ^[26,27]^ was developed to quantify the copy number of repetitive genomic elements in immune cells. For the past decade, many efforts to integrate FISH with microfluidic devices have been made to study chromosome abnormalities ^[28]^, circulating tumor cells ^[29]^, and onco-hematological malignancies ^[30]^. For bacterial identification, Liu *et al*. reported the microfluidic device-integrated Flow-FISH that enabled the detection of bacteria in environmental samples ^[31]^. Despite these extensive efforts, quantitative identification of bacteria in whole blood using FISH remains unachievable due to the complex nature of blood samples and the difficulty in enriching a broad range of bacteria cells found at a low frequency in whole blood without prior expansion.

To address this unmet challenge, here we describe a quantitative FISH approach in a microfluidic device that enables magnetic confinement of a range of bacteria in blood using opsonin-coated magnetic nanoparticles (MNPs). Our automated microfluidic-magnetic FISH (µFISH) platform can quantitatively identify bacteria in patient blood samples, which are undetectable by the conventional blood culture method within 2 hours without prior expansion *in vitro*.

## 2. Results

### 2.1. Opsonin magnetic nanoparticles and a microfluidic device for magnetically confining bacteria for FISH

All blood samples collected into DNA/RNA Shield™ blood collection tubes were preincubated with opsonin-coated MNPs to enrich bacteria contained in whole blood **(Figure 1A)** because they are often found at a low frequency in whole blood (1-10 CFU mL^-1^) ^[32,33]^. We employed human recombinant mannose-binding lectin (hrMBL) that is known to capture a broad range of bacteria, fungi, and viruses ^[34,35]^, and immobilized biotinylated hrMBL onto streptavidin-coated MNPs as described previously ^[36]^. The prepared blood samples were introduced into a microfluidic FISH device where MNP-labeled bacteria were magnetically sequestered by a permanent magnet (25 mm × 6.3 mm × 50 mm; width × height × length, BY0×04-N52, KJ Magnetics, PA, USA) **(Figure 1B)**. The microfluidic device (25 mm × 4.2 mm × 50 mm; width × height × length) consisted of two inlets (one for injecting samples and the other for flowing FISH reagent solutions) and one outlet, which was fabricated by the conventional soft-lithography method ^[37]^. We adopted a sinusoidal microfluidic channel for magnetic confinement of bacteria to increase the channel area for sequestering the bacteria **(Figure S1A)**. Multiple units of the device were connected in parallel to improve the throughput (x 8) so that we can carry out multiple FISH analyses simultaneously **(Figure S1B)**. The device was operated by a programmable pneumatic pump that controls eight pressure chambers individually (07-VB-08-100, SmartSquart®, Automate Scientific, CA, USA) so that reagents required for processing FISH were sequentially injected into the microfluidic channel as programmed **(Figure S1C)**. Reagents for fixation and permeabilization of bacteria were sequentially injected into the device, followed by incubation with FISH probes complementary to target 16S ribosomal RNA (rRNA) sequences **(Figure 1C and Figure S1D)**. After washing unbound FISH probes, fluorescence images over the entire channel area were obtained by a fluorescence microscope to quantitate bacterial concentrations in the given sample volume (**Figure 1D** and Figure S1A).

**Figure 1.**
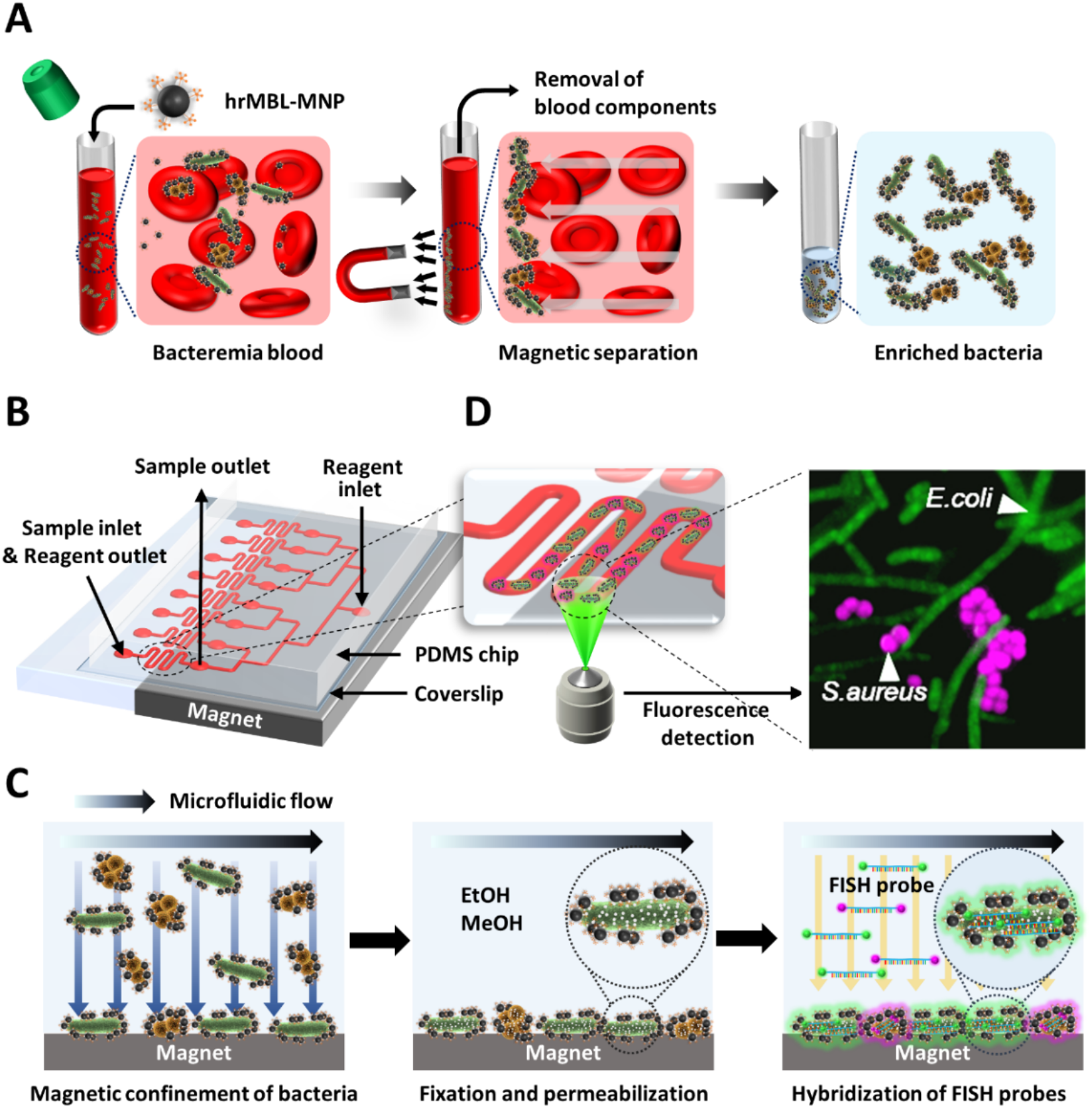
A schematic diagram of quantitative bacteremia identification using opsonin-coated MNPs and μFISH. A) Bacteria in bacteremia blood are magnetically captured and enriched by MNPs conjugated with human recombinant mannose-binding lectin (hrMBL-MNPs). B) The PDMS microfluidic device consists of eight inlets and outlets each for samples and FISH reagents, and 8 dividing channels for high throughput. C) Bacteria captured by the MNPs are magnetically confined in the microfluidic channel and then fixed and permeabilized to hybridize the fluorescence-labeled FISH probes to the complementary 16S rRNA sequences of the target microbes. D) Fluorescent imaging of magnetically captured bacteria (*E. coli*: green, *S. aureus*: magenta).

### 2.2. FISH probe design and characterization

Although FISH has been used for bacterial identification for decades, recent advances in microbial genomics bring a new challenge in designing species-specific probes. For example, the *E. coli*-specific probe (pB-1326; AGC CAT GCA GCA CCT GTC TC) ^[38]^, available in public depository probeBase ^[39]^, permits involuntary hybridization with about 10% of non-target bacterial species (1,640 out of 14,600 species according to the NCBI bacterial 16S rRNA RefSeq data). To overcome this, we designed a set of FISH probes with up-to-date microbial genome resources. Even with longer 30-bp probes targeting the conserved 16S rRNA sequences, it was unachievable to design the specific probes exclusively hybridizing with a target bacteria species among over ten thousand bacterial species with known rRNA sequences. Thus, we designed the *E. coli*- and *S. aureus*-specific probes, respectively, and evaluated that they were not at least cross-reactive to seven bacterial species commonly observed in the blood culture (*Klebsiella pneumoniae, Proteus mirabilis, Bacillus subtilis, Enterococcus faecalis, Escherichia coli, Salmonella enterica, Staphylococcus aureus*) (See the experimental section). Furthermore, we checked putative target species for these FISH probes, confirming that they were not commonly detected from the human microbiome studies (See the experimental section for the list of those bacterial species). In addition, we also set out to find the sequences of the FISH probe that is expected to bind to the vast majority of 16S rRNA of bacteria (the universal probe), which we can use for fluorescently labeling most bacterial cells. Both *E. coli*- and *S. aureus*-specific probes were labeled with Cy5 fluorophore at the 5’ end, and the universal probe was labeled with Cy3 fluorophore so that we can identify *E. coli* or *S. aureus* among a broad range of magnetically isolated bacteria by using both the species-specific (Cy5) and the universal (Cy3) probes.

The above FISH probes were tested for their detection efficiency and target specificity. For *E. coli* and *S. aureus*, both the species-specific probes and the universal probe efficiently bound to the targets **(Figure 2A, B, and Figure S2)**. The species-specific probes showed much lower signals in the other five species tested, confirming their target specificity, while the universal probe showed consistently high signals in all seven species **(Figure S3)**. The probes showed small crosstalk between the two species, 0.8% for the *E. coli*-specific probe and 0.5% for the *S. aureus-*specific probe (Figure 2B; Experimental section). A magnified view of a bacterium labeled with 4’,6-diamidino-2-phenylindole (DAPI), the universal probe (Cy3), and the *E. coli*-specific probe (Cy5) showed the distinct morphological feature of *E. coli* **(Figure 2C)**, *S. aureus*, and *K. pneumoniae* **(Figure S4A)**. We also validated the robustness of the FISH probes in the µFISH device. We quantitatively measured the averaged fluorescence intensity of *E. coli* magnetically confined in the µFISH device, which were treated with the universal probe (Cy3) and the *E. coli*-specific probe (Cy5), or with the *S. aureus*-specific probe (Cy3). The majority of *E. coli* cells were hybridized with the universal probe (99.9%) and the *E. coli-*specific probe (99.5%) but not with the *S. aureus*-specific probe (0.69%) **(Figure 2D)**.

**Figure 2.**
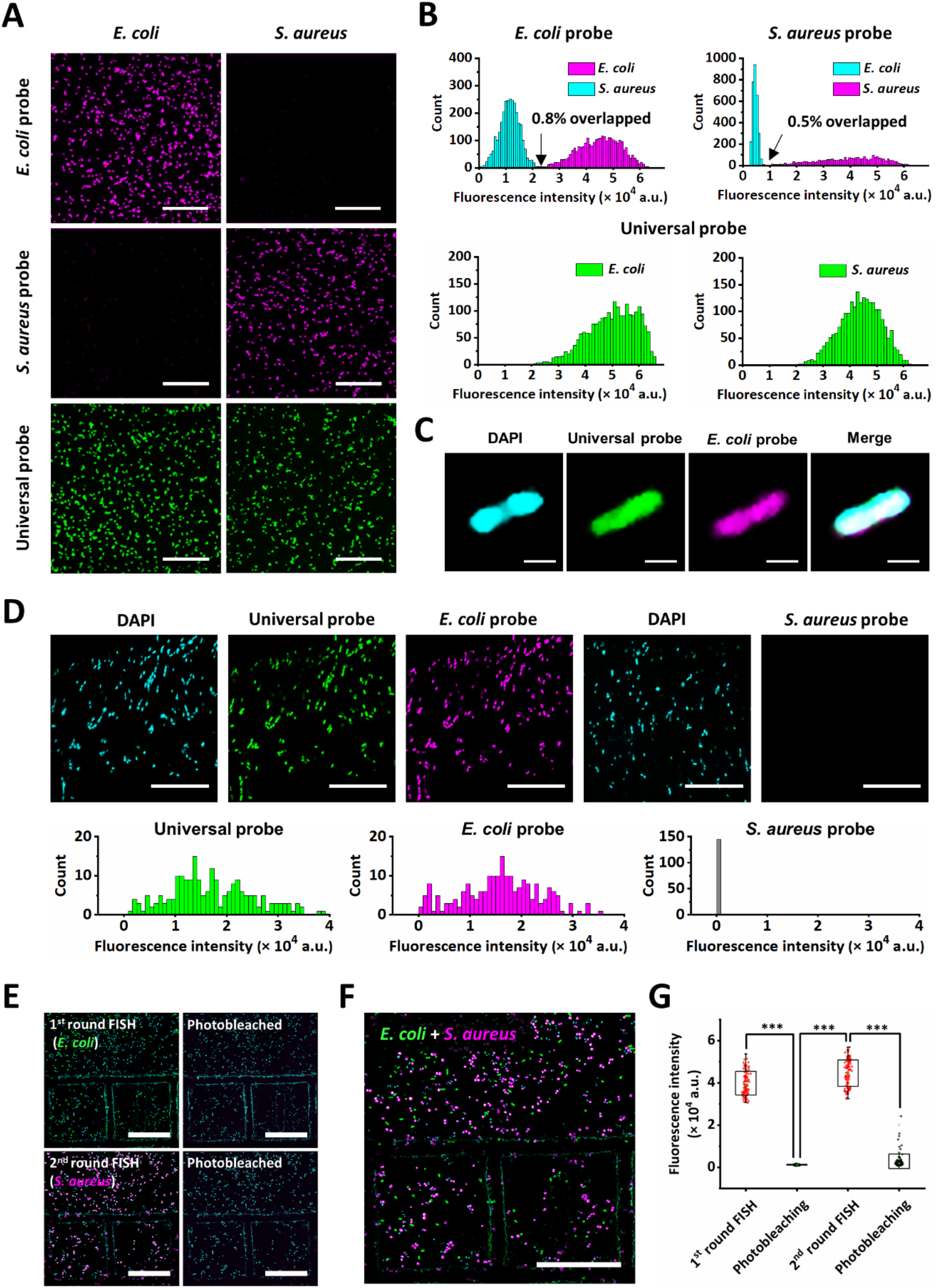
FISH probe characterization. A) Fluorescence images of *E. coli* and *S. aureus* labeled with FISH probes specific or non-specific to each of them. Scale bars, 25 µm. B) Histograms of the FISH signal intensity of 3,000 bacterial cells from each labeling combination. The FISH signal intensity of target species (magenta, green) was shown along with that of non-target species (cyan) for both kinds of the probes. Percentages represent the crosstalk across species as the overlapping portion between the populations. C) Confocal microscopy images of *E. coli* stained with DAPI, the universal bacteria probe (Cy3), and the *E. coli*-specific probe (Cy5) using a 60x objective lens. Scale bars, 1 µm. D) Fluorescence images (upper panels) emitted by DAPI and each FISH probe (universal probe and *E. coli*-specific probe), respectively, represent that the majority of *E. coli* cells magnetically captured in the device can be identified by µFISH. The average FISH signal intensity within each *E. coli* was measured to build histograms of the mean value of fluorescence intensity of each probe (graphs in lower panels). The cross-reactivity between *E. coli*-specific and *S. aureus*-specific probes were insignificant (right panel) because only 0.69% of *E. coli* non-specifically were hybridized with the *S. aureus*-specific FISH probe. Scale bars, 30 µm. E-G) Multiplexed FISH images enabled by sequential FISH and photobleaching steps. E) The first round of FISH using *E. coli*-specific probe (Cy3) visualized *E. coli* cells positioning on the grid-patterned substrate. The photobleaching using laser exposure (561 nm for 5 min) quenched Cy3-labeled probes, leaving the signals only emitted from DAPI. The secondary FISH labeled *S. aureus* with Cy3 again and then quenched by the laser irradiation. Scale bars, 10 µm. F) Overlay of images obtained from E) showing FISH signals emitted from the *E. coli*-specific probe (pseudo-colored green) and *S. aureus*-specific probe (pseudo-colored magenta). Scale bar, 10 µm. G) Scatter plot showing the fluorescence intensity distributions of bacteria at each step of consecutive FISH imaging. Bacteria were detected based on DAPI signal. (\*\*\**p* < 0.001)

We further demonstrated that the species-specific probes could exclusively detect their target species in a mixed bacteria sample. We attached an equal mixture of *E. coli* and *S. aureus* on slides and performed two rounds of sequential FISH measurements **(Figure 2E)**. In the first round, fluorescence images were taken with the *E. coli*-specific probe, and then after photobleaching with 561 nm laser irradiation at 15 mW for 5 min, in the second round, it was repeated with the *S. aureus*-specific probe. Overlaying the FISH images revealed that each probe visualized its target species with high specificity showing only one duplicate detection from 142 *E. coli* and 354 *S. aureus* cells **(Figure 2F)**. Nearly complete photobleaching of the FISH signals demonstrates that this scheme can be used for multiplexed detection of multiple bacterial species in a sample **(Figure 2G)**.

### 2.3. Quantitative bacterial FISH in a microfluidic device

It is essential to quantitate bacteria concentration directly in whole blood because no conventional diagnostic methods can provide a quantitative measure of bacterial load in whole blood even after prior blood culture. First, we measured the binding affinity of the hrMBL-MNPs to *E. coli* and *S. aureus*. Fluorescence images demonstrated that the *E. coli* and *S. aureus* (DAPI) were covered with hrMBL-MNPs labeled with Atto-550 **(Figure 3A)**, supporting that the hrMBL-MNPs can be used to capture and magnetically separate the bacterial cells we are targeting in this study **(Figure S4B)**.

**Figure 3.**
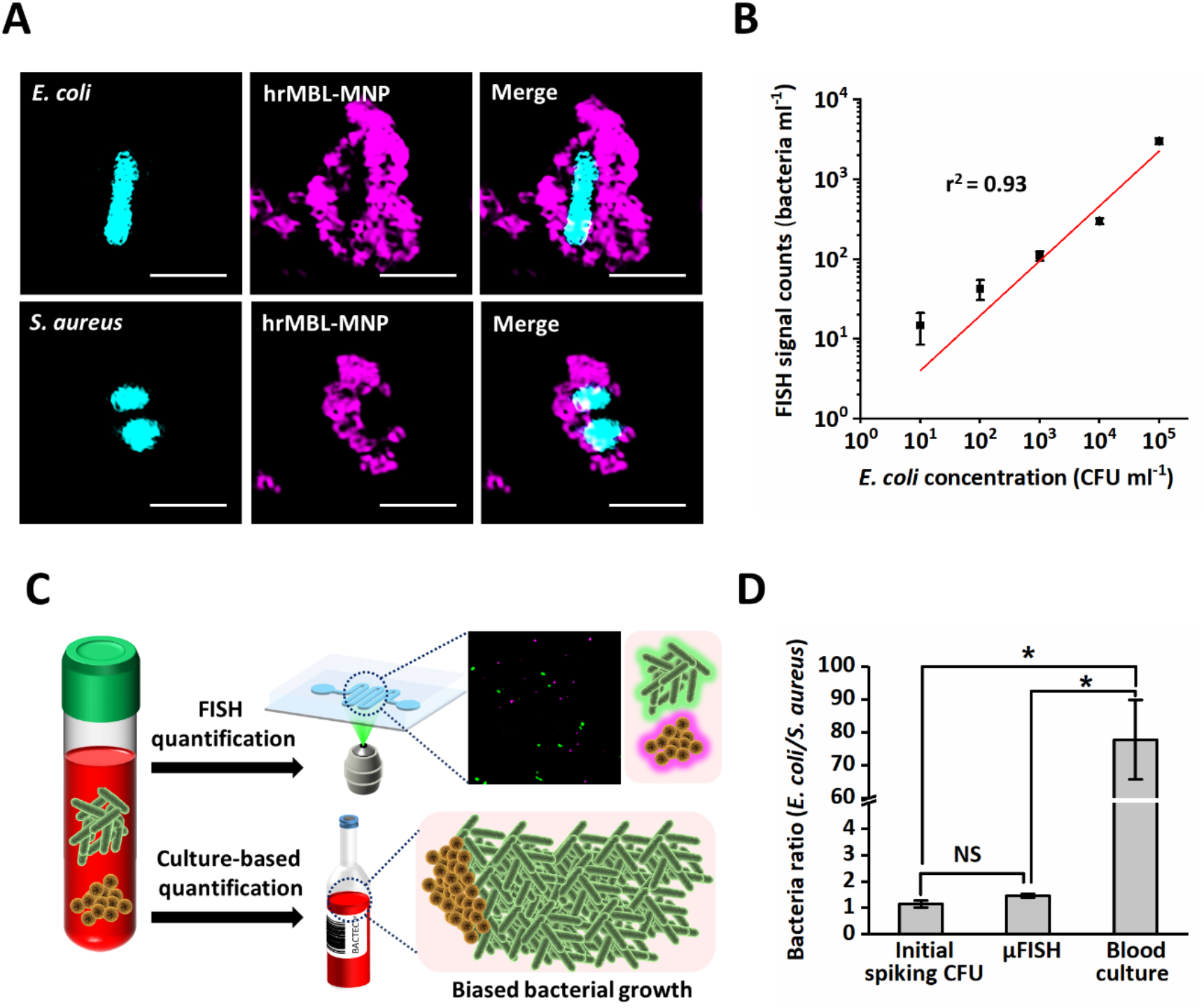
Magnetic separation of bacteria with hrMBL-MNPs and quantification of bacterial load using µFISH chip. A) Structured-illumination microscopy (SIM) images of *E. coli* and *S. aureus* (DAPI, cyan) captured by hrMBL-MNPs labeled with Atto 550 (magenta). Scale bars, 2 μm. B) Quantification of the FISH signals using every ten-fold diluted *E. coli* (10-10^5^CFU/mL) (*n* = 3 technical replicates; ± s.e.m.). All values of the FISH signals shown in the graph were subtracted by background signals, which were obtained with a sterile saline solution in the µFISH device. C) The quantitative power of µFISH in comparison to the conventional blood culture method. A schematic illustration representing that the µFISH method retains quantitative information of the initial polymicrobial composition in the sample. D) Experimental validation of the quantitative power of µFISH using a mixture of *E. coli* and *S. aureus* at a ratio (γ = *E. coli* / *S. aureus*) of 1.14 ± 0.13. γ determined by µFISH was 1.46 ± 0.07 while the conventional culture method resulted in γ of 77.7 ± 12.06 (*n* = 3 independent experiments). (NS: Not Significant, \**p* < 0.05)

Then, we set out to validate that the FISH signals proportionally correlate with bacterial concentrations in a solution. *E. coli* (K-12 W3110, Korean Collection for Type Culture (KCTC) 2223) at concentrations ranging from 0 to 10^7^ CFU mL^-1^ were prepared in a saline solution. The fluorescence signals from DAPI, Cy3 of the universal bacteria probe, and Cy5 of the *E. coli*-specific probe were measured at each concentration. The FISH signals increased linearly with the bacteria concentration ranging from 10 to 10^5^ CFU mL^-1^ and then plateaued due to a large number of the bacterial cells magnetically piled up in the sinusoidal microfluidic channel. To extend the range of the detectable bacterial concentrations above 10^5^ CFU mL^-1^, the samples containing higher *E. coli* numbers than > 10^5^ CFU mL^-1^ were diluted accordingly (x 10^−1^ or x 10^−2^) so that the final concentration of bacteria can fit within the quantitative range (10-10^5^ CFU mL^-1^) **(Figure 3B)**. Interestingly, we found that our platform can even discriminate the presence of unculturable bacteria found at a low frequency (<1-5 cells mL^-1^) in a buffer solution because we detected the triple-superimposed fluorescence signals (DAPI, Cy3, and Cy5) when using saline solutions without spiking any bacteria while all FISH reagents, magnetic particles, and buffer solutions we used were culture negative. This might be attributed to low bio-mass contamination caused by the reagents we used, the so-called “kitome” ^[40]^; however, these background signal counts were consistently low enough to be distinguished from the fluorescence signal counts of bacterial cells (Figure 3B).

Next, we validated how rapidly our µFISH system could provide quantitative information of bacteria in whole blood in comparison to the conventional diagnostic methods, including blood culture and PCR. Among the three methods, the µFISH and blood culture methods detected the presence of bacteria in rat blood spiked with *E. coli* (10^2^ CFU mL^-1^) while the PCR method was insensitive to prove their existence in blood due to the low concentration **(Table 1)**. Moreover, only our µFISH method quantitated bacterial loads in blood at the time of collection within two hours, whereas blood culture, which took 9.5 hours to obtain the positivity results, was unable to provide initial bacterial concentrations due to expansion in vitro. More importantly, we validated that our platform can reveal an initial population ratio of polymicrobial samples, which is often biased after cultivation for hours to days due to the different growth rates of diverse strains of bacteria in a given culture condition. When a mixture of *E. coli* and *S. aureus* at a ratio (γ = *E. coli* / *S. aureus*) of 1.14 ± 0.13 was spiked in rat blood, and the population ratio of two species was determined by the culture-free µFISH method and the culture-based method **(Figure 3C)**. γ measured by µFISH retained similarly (1.46 ± 0.07) while the blood culture method failed to reveal the initial population ratio (γ: 77.7 ± 12.06 (*n* = 3)) due to the different growth rates **(Figure 3D)**. This unique capability offers immense clinical impact because the conventional bacteremia diagnostic methods commonly necessitate prior blood culture, making the original population ratio mis-quantified.

**Table 1.** Comparison of µFISH with conventional bacteria detection methods (blood culture and qPCR). 10^2^ CFU mL^-1^ of *E. coli* was spiked into rat blood, and the superiority of µFISH in comparison to other methods were listed. TTP, Time-to-positivity.

| *E. coli* in blood<br>($10^2$ CFU mL<sup>-1</sup>) | Diagnosis outcomes | Time to result | Quantification |
| --- | --- | --- | --- |
| μFISH | Positive | 2 | O |
| Blood culture (TTP) | Positive | 9.5 h | X |
| qPCR | Negative | - | - |

### 2.4. Determination of bacterial species in the blood of a porcine bacteremia model

We next validated the preclinical utility of our µFISH platform using a porcine bacteremia model. The bacteremia model was developed by intra-abdominal infection using fecal contents, as described in our previous study ^[41]^, which recapitulated sepsis and bacteremia of intra-abdominal origin in human patients. Whole blood samples (3 mL) collected every two hours post-infection were processed using hrMBL-MNPs for µFISH analysis. Our previous study on the porcine bacteremia model revealed that the majority of bacteria detected by blood culture were *E. coli* over 12 h observation ^[41]^. Thus, we quantitated FISH signals by using the universal probe and the *E. coli*-specific probe. The magnetically enriched samples contained immune cells, and the activated immune cells are known to phagocytose opsonin-coated nanoparticles, such as hrMBL-MNPs ^[42]^, similar to phagocytes engulfing opsonized microbial cells. However, those immune cells did not noticeably interfere with imaging the FISH-labeled bacteria **(Figure S4C)**. We enumerated the number of bacteria or *E. coli* by superimposing the images of DAPI and the universal probe (Cy3) or DAPI and the *E. coli*-specific probe (Cy5), respectively.

We found that the porcine blood samples collected even before inducing fecal peritonitis in the intra-abdominal cavity consistently retained unculturable bacteria in a range of 10^2^-10^3^ bacteria mL^-1^ of blood, and the bacterial levels changed in the blood throughout 12 h of fecal peritonitis. It should be noted that the microfluidic FISH device quantitatively determined the presence of bacteria and *E. coli* in blood for the entire time points, while the conventional blood culture methods detected them only after inducing fecal peritonitis. Although we do not have clear evidence underpinning the unculturable bacteria in the porcine blood, we suspect that antibiotics, which were already given to the pigs on the farm, might have inactivated pre-existing bacteria in the blood. This was substantiated by the detection of endotoxin in the blood even before inducing fecal peritonitis in our porcine bacteremia model **(Figure S5)** ^[41]^. However, the µFISH device was able to magnetically enrich rare bacteria (viable or non-viable) and their debris in a confined area of the microfluidic channel and optically identify and enumerate them simultaneously **(Figure 4A)**. As predicted by the blood culture results, *E. coli* population takes up the majority of bacteria in the blood samples as quantitated by the µFISH device. The bacterial levels in whole blood of the porcine model were not consistent over the time course for all pigs we implemented **(Figure 4B-F)** because of the complexity of the pathogenesis of peritonitis-induced bacteremia and the immune status of each pig. We obtained blood culture positive results (*Staphylococcus* species) from two pigs before inducing fecal peritonitis (T0 of Figure 4C, F), which is most likely due to contamination that often occurs at the skin ^[43]^. Most importantly, the bacterial load determined at each time point by the µFISH device closely correlated with the hemodynamic variables, such as stroke volume and cardiac index measured at each time point, and the *E. coli* µFISH signals also corresponded to the endotoxin levels in pig blood (*p* < 0.05). **(Figure 4G)**. This implies that the bacterial quantification by µFISH could offer the predictive power of the clinical indices when the platform becomes available for patients.

**Figure 4.**
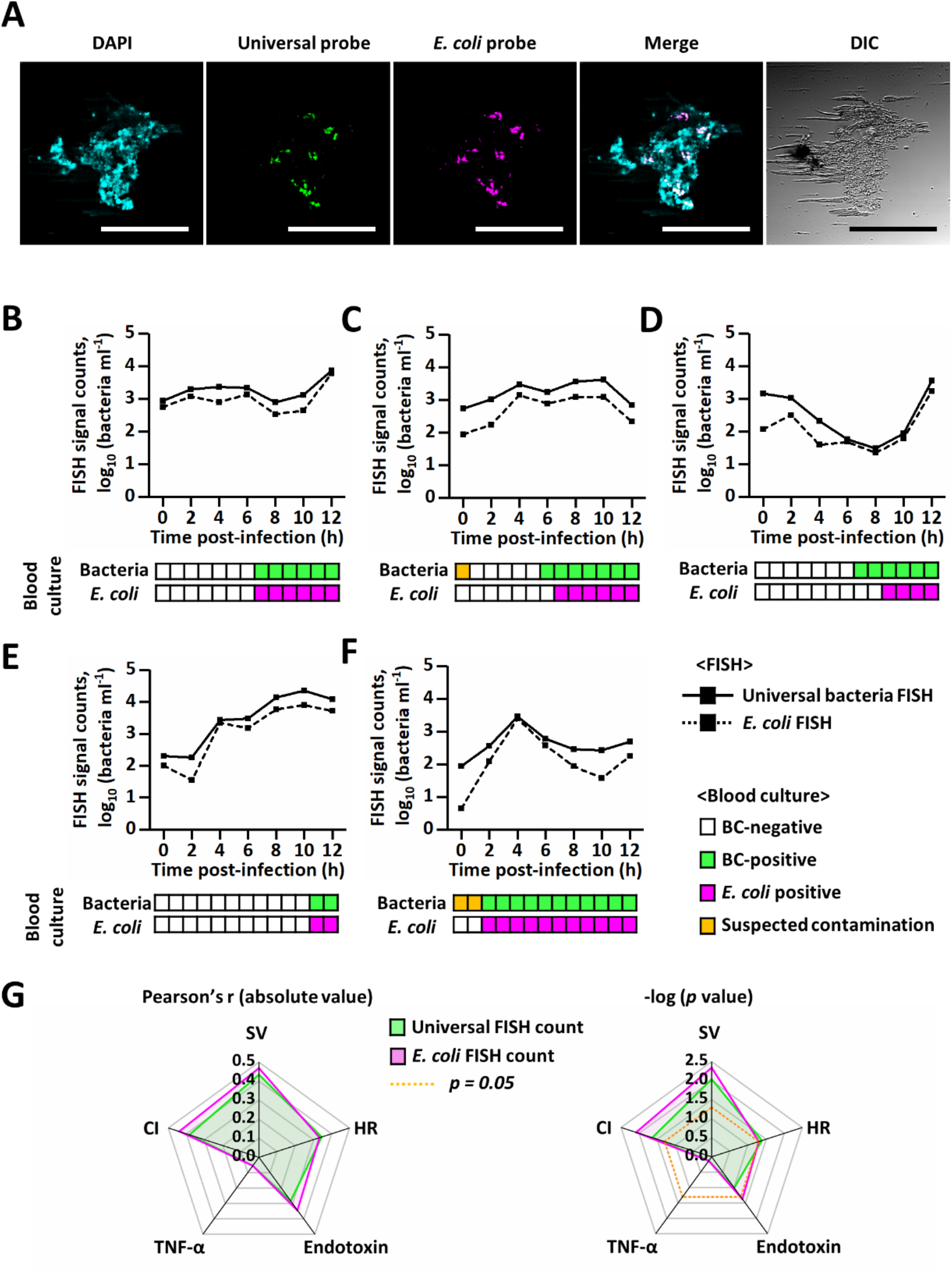
Quantitative identification of bacteria in porcine bacteremia blood using µFISH. A) *E. coli* from porcine blood samples were determined by the FISH images superimposed with DAPI, Cy3, and Cy5 signals. Cyan: DAPI, green: universal bacteria probe-Cy3, magenta: *E. coli*-specific probe-Cy5. Scale bars, 40 μm. B-F) The quantitative FISH signal counts of the blood samples of the five individual porcine bacteremia models over the time course of infection up to 12 hours (*n* = 5 individual porcine models). µFISH revealed bacteria and *E. coli* in bacteremia blood at the earlier time points than the conventional blood culture method. In addition to reporting the presence of the bacterial cells in blood, µFISH enabled a quantitative measure of both overall bacterial loads and the particular species in biological samples. G) Polar charts showing the correlation between FISH signal counts and physiological parameters of the porcine bacteremia model analyzed by Pearson’s correlation coefficient *r* values (absolute). Statistical significance was verified with *p*-values from one-way analysis of variance (ANOVA). Green: Universal FISH counts, Magenta: *E. coli* FISH counts. CI, Cardiac index; SV, Stroke volume; HR, Heart rate.

### 2.5. Pilot clinical validation of the FISH device with human patient blood samples

We next validated the clinical utility of the µFISH device using patient blood samples. We carried out a retrospective clinical study using the blood samples collected from the suspected sepsis patients and stored until testing **(Table S1)**. The blood samples had been collected prospectively in adult patients with suspected sepsis who visited the emergency department (fever and quick sequential organ failure assessment score ≥ 2). 3 mL of each blood sample was taken into DNA/RNA Shield™ blood collection tubes before antibiotics administration. Blood samples of ten patients who were confirmed with *E. coli* infection afterward by the culture study of blood or body fluids collected from the primary sites of infection were selected. The primary sites of infection included the urinary tract (seven patients), the biliary tract (two patients), and the genital tract (one patient). The culture study of body fluids isolated from all enrolled patients’ primary sites of infection revealed *E. coli* as a causative pathogen. Of these ten patients, No. 1, 6, and 8 patients (30%) had *E. coli* bacteremia confirmed by the conventional blood culture while No. 10 patient had a different kind of bacteria in the blood (*Klebsiella pneumoniae*), and the other six patients (No. 2, 3, 4, 5, 7, and 9) were blood culture-negative. We then quantitated the bacterial and *E. coli* levels in blood samples of all ten patients using our µFISH device **(Figure 5A)**. While human blood without infection showed the background levels of the µFISH signals **(Figure 5B)**, the quantitative µFISH results revealed that *E. coli* were present in the blood samples of all ten patients (sensitivity: 100%, **Figure 5C**). In addition, we corroborated that a ratio of *E. coli* counts to total bacterial loads in each sample measured by quantitative µFISH signals correlated with that calculated by normalizing read counts of bacteria and *Escherichia*/*Shigella* genus in the patient blood samples from microbiome profile experiment (Experimental section) (*p* = 0.0014) **(Figure 5D, E)**. This supports the quantitative capability of µFISH that allows to not only identify bacteria in biological samples but quantitate bacterial loads without next-generation sequencing (NGS) analysis.

**Figure 5.**
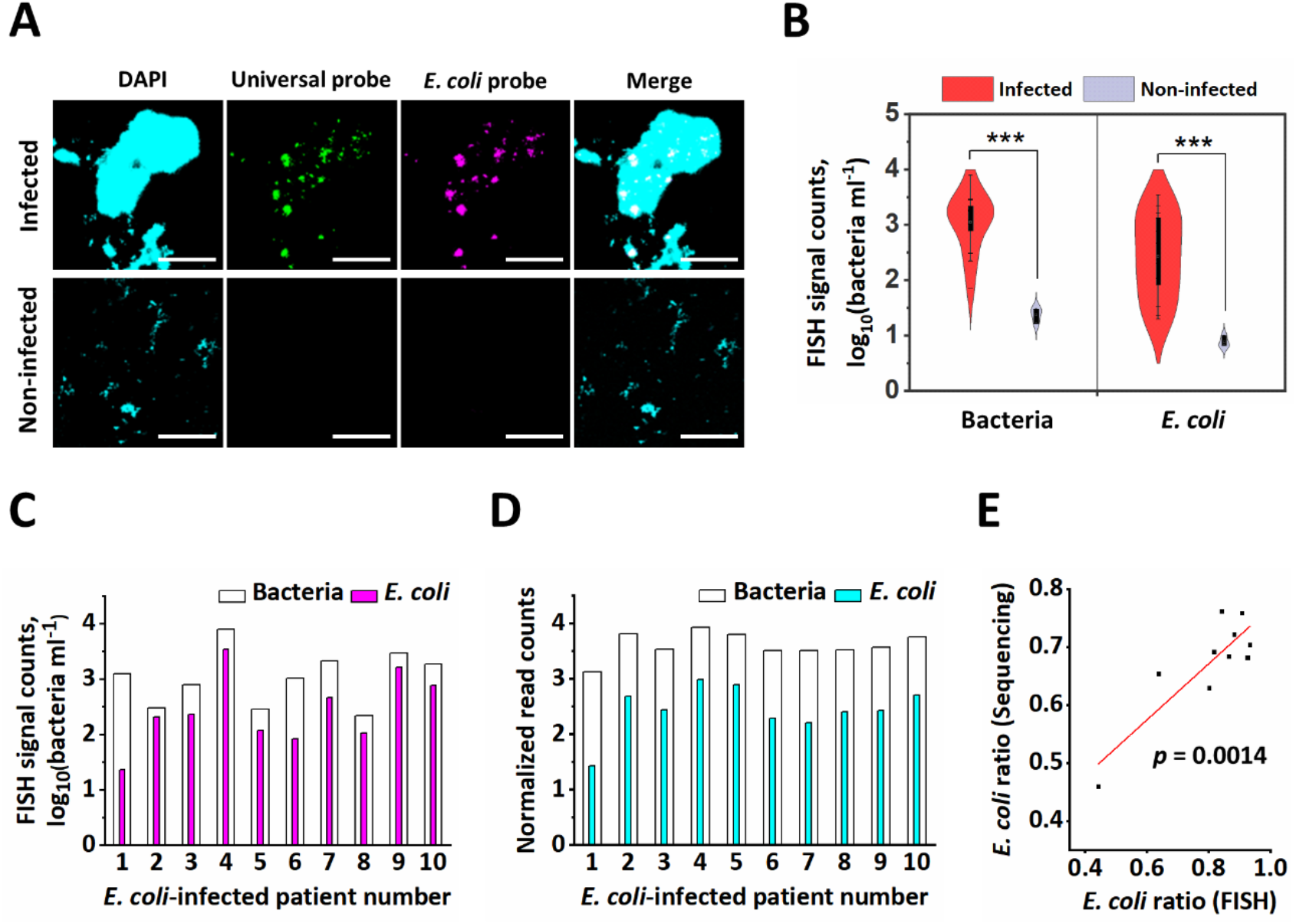
Quantitative identification of *E. coli* in blood samples of patients with infection using µFISH and sequencing. A) Fluorescent images of *E. coli* labeled with the universal probe (green) and the *E. coli*-specific probe (magenta) in blood samples from the *E. coli-*infected patients and donors without infection, respectively. Cyan: DAPI, green: universal bacteria probe-Cy3, magenta: *E. coli*-specific probe-Cy5. Scale bars, 50 µm. B) Universal bacterial loads (left) and *E. coli* levels (right) in whole blood of the patients with *E. coli* infection were determined by µFISH. Blood samples collected from human donors without infection (*n* = 3) resulted in low background levels of FISH signals, whereas those of *E. coli*-infected patients (*n* = 13) presented distinguishable levels of microbes and *E. coli* in blood. (\*\*\**p* < 0.001) C-D) Quantitative profiles of the microbiome in blood collected from patients infected with *E. coli* (*n* = 10) measured by µFISH (C) and Illumina sequencing (D). E) A correlation between a ratio of *E. coli* to total bacterial loads in each patient’s sample measured by µFISH and sequencing was calculated (*p* = 0.0014). In µFISH, *E. coli* counts (*E. coli*-specific probe) were normalized to total bacteria counts (universal probe). For the sequencing method, we normalized the total number of the reads and those mapped to the *Escherichia*/*Shigella* genus to the read counts corresponding to the three spike-in species (*Truepera, Imtechella*, and *Allobacillus*) for absolute quantification. Those three spike-in species are known to be not found in humans.

## 3. Discussion and Conclusion

Identification and quantification of both viable and non-viable bacteria in patient blood have been elusive due to the lack of capable tools despite their clinical importance, particularly for immunologically vulnerable patients. We demonstrated the versatility of the FISH technique combined with the opsonin-coated MNPs and the automated microfluidic control system. The clinical validation using patient blood samples evidenced its capability of determining causative pathogens even in the blood culture-negative cases.

In the porcine bacteremia model, we first identified a significant correlation between the quantitative bacterial load measured by the µFISH device with hemodynamic parameters, such as stroke volume and cardiac index, which had been unachievable because of the lack of tools to quantitate bacterial levels in whole blood. Our device also detected bacteria in pig blood even before inducing intra-abdominal peritonitis, which is most likely due to dormant blood microbiome possibly translocated from their gut ^[44]^.

Our system determined the presence of *E. coli* in the blood of the patients who had *E. coli* as the common causative pathogen in each infection site regardless of the blood culture positivity. Because the bacterial load in blood is often associated with clinical outcomes in patients with bacteremia ^[5,45,46]^, we set out to extrapolate the correlation between the severity of the septic patients with a greater number of FISH signal counts. Although the FISH signal levels did not correlate with the patients’ severity, most likely because the majority of bacteria detected by the µFISH device were not culturable, a more insightful explanation could be made after follow-up clinical studies with a large number of enrolled patients in the future. Moreover, in patient No. 10, the culture of bile juice directly acquired from the gallbladder puncture revealed the causative pathogens of *E. coli* and *K. pneumoniae*, while the majority of FISH signals of the patient blood was identified as *E. coli*, which could be attributed to the difference in the survival of the strains in human whole blood ^[47,48]^ and variable growth rates in a subsequent culture condition.

Due to the extensive diversity of bacterial species and a limited set of distinguishable fluorophores, it is impossible to identify all bacterial species in one round of µFISH. Employing the multiplexed FISH capability we demonstrated above, we can rapidly narrow down to potential causative microbe candidates by carrying out multiple µFISH measurements using different sets of probes sequentially. For example, we may determine loads of Gram-positive or Gram-negative bacteria in the first round of µFISH, followed by rounds of µFISH for more precise taxonomic identification. In a clinical setting, it is critical to gain instantaneous information on whether causative bacteria are Gram-positive or Gram-negative because it determines the types of antibiotics to be given to the patients.

In contrast to the conventional bacterial identification methods, such as PCR and mass spectrometry, our approach allows us to quantitatively determine the presence of intact bacteria in collected samples, which would be clinically more impactful than validating the existence of DNA or RNA fragments from bacteria using PCR or NGS. More extensive microbiome studies could enable probe design with higher specificity because the hybridization capability of the FISH probe depends on genomic resources available in the public depository. To improve the specificity and sensitivity of our platform, FISH probes labeled with multiple fluorophores could be employed, as in multi-color FISH techniques ^[49]^, so that we can identify multiple bacterial species simultaneously in a single round of µFISH. Combining the sequential µFISH capability we presented with the multi-color FISH probes, we could extend the boundary of µFISH technique further to studying microbiome residing on various epithelium surfaces, which could provide not only taxonomic information but also their spatiotemporal distribution in the microenvironment.

## 4. Experimental Section

### Preparation of opsonin-bound magnetic nanoparticles

For targeting a wide range of bacteria, human recombinant mannose-binding lectin (hrMBL) (10405-HNAS, Sino Biological, China) was conjugated with MNPs in 200 nm size ^[50]^. Before the conjugation, hrMBL (0.25 mg mL^-1^) was biotinylated using 10 mM biotinylation reagent (A39256, Thermofisher, MA, USA) for 30 min at room temperature. Then, streptavidin-coated MNPs (200 nm in diameter; 03121, Ademtech, France) were mixed with the biotinylated hrMBL for 30 min and washed with 1X phosphate-buffered saline (PBS) supplemented with 1% bovine serum albumin. The conjugated hrMBL-bound MNPs were stored at 4 °C before use.

### Microfluidic device fabrication and operation

To construct microfluidic devices for bacteremia detection, a serpentine microchannel with a height of ∼200 μm was designed, and its silicon wafer mold was fabricated by standard photolithography with SU-8 photoresist (SU-8 2150, MicroChem Corp., MA, USA). The wafer was then silanized overnight before replicating Polydimethylsiloxane (PDMS) (Sylgard 184 A/B, Dow Corning, MI, USA) microchannel devices. Heat-cured PDMS devices were UV-sterilized and bonded onto UV-sterilized coverslips by plasma treatment followed by heat curing. The prepared devices were sequentially treated with 70% ethanol, DEPC-treated water, and then primed with 2% Pluronic F108 (542342, Sigma-Aldrich, MO, USA) solution for 2 hours to block unnecessary molecules from non-specific binding on the surface of the devices and washed with 1X TBST buffer (TR2006-100-00, Biosesang, South Korea) with 5 mM CaCl2 (21115, Sigma-Aldrich) until use. Then, samples were injected into the devices using a syringe pump (Fusion 200 Touch, Chemyx Inc., TX, USA) and magnetically pulled by locating a magnet (BYO88-N52, NdFeB, grade N52, KJ Magnetics, PA, USA) beneath (**Figure S1B**, left panel). After magnetically capturing the bacteria in the sinusoidal channel, FISH reagents were sequentially introduced into the device to carry out µFISH on the captured bacterial cells (Figure S1B, right panel).

### FISH probe design

We used the NCBI bacterial 16S ribosomal RNA RefSeq data (BioProject accession number PRJNA33175, downloaded on June 14th, 2019) to design the probes for FISH. Out of 20,160 sequences, we excluded the sequences longer than 2,000 bp or those shorter than 1,000 bp, and 19,885 sequences from 14,600 bacterial species were retained. We extracted all 30 bp oligonucleotide sequences from this database and searched their putative matched targets using EXONERATE^[51]^, with ungapped local alignment mode allowing 2 bp mismatches. Because it is unachievable to find a probe sequence exclusively mapped on a single species when we consider all available bacterial 16S rRNAs, we only included seven bacterial species (*Klebsiella pneumoniae, Proteus mirabilis, Bacillus subtilis, Enterococcus faecalis, Escherichia coli, Salmonella enterica, Staphylococcus aureus*) commonly found in the blood culture of human patients with bacteremia as a target to prevent cross-hybridization ^[52]^. As a result, *E. coli*- and *S. aureus-*specific 30 bp FISH probes were designed, as shown in **Table 2**. A universal probe targeting a highly conserved region of 16S rRNA of a broad range of bacteria was designed based on the oligonucleotide frequency of the mainly targeted seven bacterial species and its length was manually trimmed to improve the coverage. The universal probe used in this study (Table 2) was designed to target 10,518 species (10,632 species with an allowance of 2 bp mismatch) in the reference database used for the probe design, which covers 72% of all known bacterial species. The probe sequence information and fluorescent dye labeling are described as below.

**Table 2.**
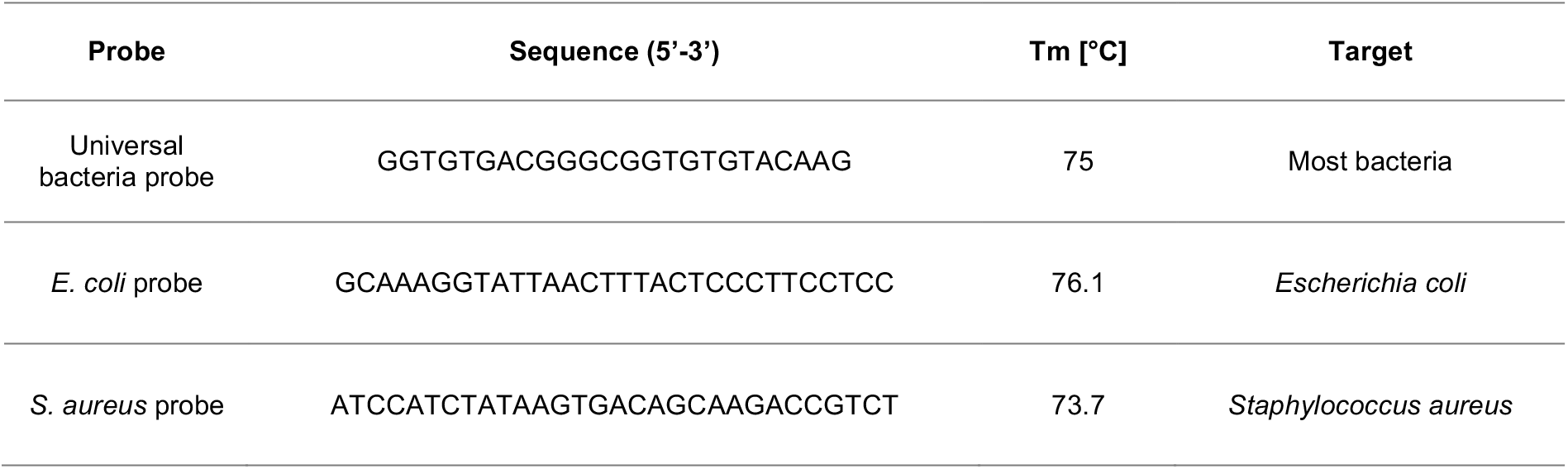
DNA FISH probes for universal bacteria (24 bp), *E. coli* (30 bp), and *S. aureus* (30 bp) were designed. Tm, Melting temperature.

- Universal probe: 5’-Cy3-GGTGTGACGGGCGGTGTGTACAAG-3’

*-E*.*coli*-specific probe: 5’-Cy5-GCAAAGGTATTAACTTTACTCCCTTCCTCC-3’

- Other targets (10): *Brenneria alni, Escherichia albertii, Escherichia fergusonii, Escherichia marmotae, Pectobacterium carotovorum, Pseudescherichia vulneris, Shigella boydii, Shigella dysenteriae, Shigella flexneri, Shigella sonnei*

*- S. aureus*-specific probe: 5’-Cy5-ATCCATCTATAAGTGACAGCAAGACCGTCT-3’

- Other targets (9): *Staphylococcus aureus, Staphylococcus capitis, Staphylococcus caprae, Staphylococcus devriesei, Staphylococcus epidermidis, Staphylococcus pasteuri, Staphylococcus saccharolyticus, Staphylococcus simiae, Staphylococcus warneri*

5’-C6-amino modified DNA probes (Custom DNA Oligos, Mbiotech, Korea) were labeled with fluorescent dyes (Cy3 NHS ester/Cy5 NHS ester, Lumiprobe, MD, USA). For labeling reaction, the DNA probes (44 µM) and the fluorescent dye (1.54 mM) were mixed in 0.1 M sodium tetraborate decahydrate buffer with gently shaking in dark for 6 hours at room temperature, followed by DNA precipitation using ethanol according to the generalized protocol. Finally, fluorescence labeling efficiency of the DNA FISH probes was confirmed by NanoDrop 2000 (Thermofisher, MA, USA) and stored at −20 °C with minimum light exposure before use. The efficiency of fluorescent labeling was above 75% for all oligos.

### Bacterial identification using qPCR and blood culture

Rat blood samples were acquired from eight to ten-week-old male Wistar rats (Orient Bio Inc., Korea) following the animal ethical guidelines approved by the Institutional Animal Care and Use Committee (IACUC) of Ulsan National Institute of Science and Technology (UNIST) (approval number: UNISTIACUC-18-03). For a blood sample spiked with *E. coli* (10^2^ CFU mL^-1^), 3 vials of *E. coli* Bioball® (∼90 CFU) (413833, NCTC 12923, SingleShot Bioball^®^, BioMérieux, France) were added into 0.9 mL of anti-coagulated rat blood. For a polymicobial blood sample, each one vial of *E. coli* Bioball (∼30 CFU) (413833, NCTC 12923, SingleShot Bioball™, BioMérieux, France) and *S. aureus* Bioball (∼30 CFU) (56045, NCTC 10788, SingleShot Bioball™, BioMérieux, France) were spiked into 1 mL anti-coagulated rat blood. Then, the identification and quantification of the bacteria-spiked blood samples were conducted using qPCR and the blood culture method. For qPCR, DNA was extracted from the blood samples using the QIAamp DNA mini kit (51304, Qiagen, Germany) according to the manufacturer’s manual, and the eluted DNA was subjected to a qPCR reaction as a template. Primer sequences for *uidA* (*E. coli*) *and nuc* (*S. aureus*) gene amplification used in the study were as follows: *uidA* (QPK-101, TaqMan™ assay, TOYOBO, Japan) (FW: CAACGAACTGAA CTGGC AGA; REV: CATTACGCTGCGATGGAT; Probe: CCCGCCGGGAATGGTGATTAC), *nuc* (QPK-201, SYBR^®^ Green assay, TOYOBO, Japan) (FW: GACCTTTGTCAAACTCGACTTCA; REV: ACACCTGAAACAAAGCATCCTAA).

The qRT-PCR reaction was run using CFX Connect™ Real-Time System (BIO-RAD, CA, USA) under the following conditions: *uidA* : 3 min at 50 °C, 10 min at 95 °C, followed by 45 cycles of 10 s at 95 °C, 10 s at 60 °C, and 10 s at 72 °C; *nuc*: 2 min at 50 °C, 10 min at 95 °C, followed by 45 cycles of 15 s at 95 °C, 1 min at 60 °C. Quantitative data analysis was carried out by CFX Maestro 1.1 software (BIO-RAD). For the blood culture method, the blood samples spiked with *E. coli* (10^2^ CFU mL^-1^) were aseptically introduced into blood culture bottles (BD BACTEC™ aerobic medium, BD, NJ, USA), transferred to a blood culture system (BD BACTEC™ FX, BD, NJ, USA), and finally examined for time-to-positivity. To quantify the polymicrobial blood sample, the samples were introduced into blood culture bottles (BD BACTEC™ aerobic medium, BD, NJ, USA), and incubated for 5 days at 37 °C in a shaking incubator (200 rpm). Then, the BACTEC solutions were withdrawn using sterile syringes and processed for qPCR quantification as described above.

### Porcine bacteremia model

Animal experiments using pigs to induce bacteremia were approved by the IACUC in Seoul National University Bundang Hospital (approval number: BA1804-246/040-01). The pigs were handled and cared for according to the guide for the care and use of laboratory animals of the National Institutes of Health. The detailed experimental procedures were described previously ^[41]^. Briefly, autologous feces (1 g kg^-1^), which were collected a day before each experiment and diluted with dextrose saline (10 g dL^-1^) were introduced into the peritoneal cavity through a small midline celiotomy. After inducing the peritonitis, five pigs were resuscitated and monitored for 12 hours by maintaining their mean arterial pressures over 65 mmHg using maintenance fluids and vasopressors as appropriate. 3 mL of blood samples were collected into DNA/RNA Shield™ blood collection tubes (R1150, Zymo Research, CA, USA) at baseline and every 2 hours for FISH analysis, and additional 10 mL of blood samples were collected into blood culture bottles (BD BACTEC™ aerobic/anaerobic medium, BD, NJ, USA) at each time point for carrying out the blood culture concurrently. For endotoxin and cytokine assay, plasma samples were obtained by centrifugation for 15 min at 3000 rpm using a refrigerated centrifuge and stored at −70 °C until testing. The stroke volume (SV) and cardiac index (CI) were measured using a pulmonary artery catheter (Swan-Ganz catheter, model 131HF, 7 Fr; Edwards Lifesciences, CA, USA). TNF-α levels in blood were measured from the stored plasma samples using an enzyme-linked immunosorbent assay kit (PTA00, R&D Systems Inc., MN, USA) according to the manufacturer’s instruction, and the Pierce™ LAL Chromogenic Endotoxin Quantitation kit (88282, Thermofisher, MA, USA) was used for measuring endotoxin levels.

### Clinical samples

This study was conducted in accordance with Helsinki Declaration and ICH-GCP, and the institutional review board (IRB) of Seoul National University Bundang Hospital approved the prospective collection of blood samples from sepsis patients (approval number: B-1812-511-304). Patients were informed of the study, and the written consents were obtained from the patients or the legal guardians. Febrile patients who were admitted to the emergency department with suspected sepsis were screened by quick Sequential Organ Failure Assessment (qSOFA) score ≥ 2, and the SOFA score was calculated using a routine laboratory test thereafter. Acute physiologic and chronic health evaluation (APACHE 2) score was calculated using the worst values for the initial 24 hours after hospitalization. Blood samples of the patients were examined for the blood culture and additional blood samples (3 mL) were collected into DNA/RNA Shield™ blood collection tubes (R1150, Zymo Research, CA, USA) for FISH analysis. Human blood without infection was obtained from the Korea Red Cross after receiving the approval reviewed by the IRB of Ulsan National Institute of Science and Technology (UNISTIRB-16-30-G and UNISTIRB-19-23-C).

### Processing of blood samples using magnetic concentration

Blood samples (3 mL) of the porcine bacteremia model and septic patients were prepared in the DNA/RNA Shield™ blood collection tubes (R1150, Zymo Research, CA, USA). Then, hrMBL-coated MNPs (2 mg mL^-1^) were added into the samples supplemented with 5 mM CaCl2, and steady agitation was conducted by an overhead shaker for 20 min at room temperature. The samples were then exposed to magnetic fields formed by a Halbach array of N52 magnets (BYO88-N52, KJ Magnetics, PA, USA) for selectively concentrating bacteria captured by hrMBL-MNPs and washing away unnecessary blood components. The concentrated samples were stored in the DNA/RNA Shield™ solution (R1200, Zymo Research, CA, USA) at −20 °C before use. All processes above were conducted in an aseptic experimental setting.

### FISH procedures

FISH probes were evaluated using *E. coli* (K-12 W3110, KCTC 2223) and *S. aureus* (NCTC 10788) strains. Bacteria were grown to OD600 = 0.4-0.6, followed by 10-fold concentration. 10 µL of the bacteria sample was attached on a poly-l-lysine coated confocal dish (101350, SPL Life Sciences, Korea) for 30 min at room temperature. Bacteria were fixed by 3.7% (v/v) formaldehyde in PBS for 30 min at room temperature, then permeabilized by 5 mg mL^-1^ lysozyme (L1013, Biosesang, Korea) for 30 min at 37 °C and 70% (v/v) ethanol (4023-2304, Daejung, Korea) for 2 hours at 4 °C. Hybridization solution^[53]^ was prepared by mixing 15% (w/v) ethylene carbonate (E26258, Sigma-Aldrich, USA), 20% (w/v) dextran sulfate (D8906, Sigma-Aldrich, USA), 600 mM NaCl (AM9759, Invitrogen, CA, USA), 10 mM citrate buffer (C2488, Sigma-Aldrich, USA), finally adjusted to pH 6.2, and stored at −20 °C before use. The fixed bacteria were hybridized with the 100 μL hybridization solution containing with fluorophore-labeled DNA probes (1 µM) for 18 hours at 45 °C in a moist chamber. After the hybridization, unbound probes were washed out using 200 µL of 2X saline-sodium citrate (SSC) (AM9763, Invitrogen, USA) with 40% (v/v) formamide (F9037, Sigma-Aldrich, USA) twice for 45 min each at 30 °C. The samples were DAPI stained for 30 min and washed with 2X SSC buffer before imaging.

FISH in the microfluidic devices was performed by labeling magnetically captured bacteria samples inside the microfluidic devices with a continuous flow (10 μL min^-1^) of FISH reagents driven by a pneumatic control pump. For fixation and permeabilization, the samples were treated with 24% (v/v) ethanol in 1X TBST with 5 mM CaCl2 (5 min), 1X TBST with 5 mM CaCl2 (3 min for washing), and 99% methanol (5 min) (67-56-1, Biosesang, Korea) sequentially. Next, the fixed samples were incubated with the hybridization solution containing FISH probes (0.5 µM) at 45 °C for 30 min to 2 hours, followed by the same washing step and DAPI staining step as described above.

### Sequential FISH

Each bacterium (*E. coli* and *S. aureus*) was grown to 0.4-0.6 of OD600, followed by 10-fold concentration in PBS buffer. A Vectabond-coated confocal dish with grids (81148, ibidi GmbH, Germany) was incubated with the 10 µL sample for 30 min at room temperature. Bacteria were fixed by 3.7% (v/v) formaldehyde in PBS for 30 min at room temperature, and then permeabilized by 5 mg mL^-1^ lysozyme (L1013, Biosesang, Korea) for 30 min at 37 °C and by 70% (v/v) ethanol (4023-2304, Daejung, Korea) for 2 hours at 4 °C. For carrying out the first round of FISH, the bacteria mixture was hybridized with 100 µL hybridization solution pre-mixed with 1 µM of *E. coli*-specific FISH probes (Cy3) for 12 hours at 45 °C in a moist chamber. After washing unbound probes with 2X SSC with 40% (v/v) formamide twice for 45 min each at 30 °C, the samples were stained with DAPI for 30 min, washed with 2X SSC, and imaged with a fluorescence microscope.

Photobleaching was achieved by exposing a 561 nm laser (15 mW at the sample location) for 5 min to remove FISH signals of a region of interest. Then, the second round of FISH for visualizing *S. aureus* was conducted with the same experimental protocol as the first round of FISH except for the use of the *S. aureus*-specific FISH probe instead of the *E. coli*-specific FISH probe. Colocalization of the fluorescence images was achieved by aligning the secondary FISH image to the first-round FISH image by geometric transformation, calculated by phase correlation between their bright-field images ^[54]^.

### Image acquisition

Images were acquired by a Zeiss LSM 780 Configuration 16 NLO multi-photon confocal microscope (Zeiss, Germany) with DAPI, Cy3, Cy5 fluorescence filter sets. 405 nm, 561 nm, and 633 nm excitations were used for the DAPI, Cy3, and Cy5 channels, respectively. A whole image view of the serpentine microchannel of the device was taken by using a tile scanning function of the microscope for FISH signal detection. For the evaluation of FISH probes, the edges of bacteria were found from the DAPI channel image binarized with an intensity threshold that maximizes the number of detected bacterial cells.

The average FISH signal intensity within each detected bacterium was measured to build histograms showing 3,000 bacterial cells each. The crosstalk for each species-specific probe was defined as the ratio of the overlapping area between the histograms obtained for its target species and non-target species, respectively. For bacteria detection in the microfluidic device, the size (0.5-5 μm) and the merged fluorescence signals (DAPI + Cy3, DAPI + Cy5, and DAPI + Cy3 + Cy5) were considered for determination of the presence of bacteria and used for quantification of bacteria in the samples using ImageJ (NIH, MD, USA).

### 16S rRNA profiling of patient blood samples

\10 µL of the ZymoBIOMICS Spike-in Control II (D6321, Zymo Research, CA, USA) was added to the magnetically enriched blood samples as a bacterial spike-in for achieving absolute quantification. After incubating with 10 mg mL^-1^ of lysozyme (10837059001, Roche, Switzerland) diluted with 10 mM of Tris-HCl (pH 8.0) at 37 °C for one hour, the samples were incubated with lysis buffer solution (10 mM of Tris-HCl (pH 7.4), 10 mM of EDTA, and 2% of SDS) supplemented with 0.5 mg mL^-1^ of proteinase K (B-2008, GeNetBio, Korea) at 37 °C overnight. Then, genomic DNA was extracted by treating a Phenol/Chloroform/Isoamyl Alcohol mixture (25:24:1, v/v) (PC2026-050-80, Biosesang, Korea), and then incubating with 60 % volume of isopropanol and 10 % volume of 3 M sodium acetate at −20 °C overnight. After washing the pellet with 70 % ethanol, RNase A (B-2007, GeNetBio, Korea) was treated at 37 °C for 30 min, and the sample was collected with the Zymo DNA Clean & Concentrator-5 (D4014, Zymo Research, CA, USA) for DNA purification. For the library construction, we followed the conventional Illumina V34 sequencing protocol for a microbiome study and sequenced the library with Illumina iSeq 100 (Illumina, CA, USA) with a 2 × 150 bp configuration. We assigned the taxonomy information for each paired reads using the RDP classifier (version 2.11) and the database (release 11.5) with a confidence score higher than 0.8 ^[55]^. To define the operational taxonomic units (OTUs), V34 amplicons were clustered using VSEARCH (version 2.13.6) with 99% identity. The ambiguous clusters were removed and reads were included if they contain more than 5% of reads assigned to a different genus compared to the seed genus assigned in the sample cluster. The raw read counts were normalized based on the sum of reads assigned to three spike-in species, and the normalized reads assigned to the *Escherichia*/*Shigella* genus out of the total number of reads were compared with the data we obtained from the FISH experiment.

### Statistical analysis

Statistical significance of all experimental values was verified with the two-tailed Student’s *t*-test and presented as a mean of at least triplicate samples with a standard error of the mean (s.e.m.). The statistical correlation was determined by Pearson’s correlation analysis (*p* < 0.05). The OriginPro 2020 software (OriginLab, MA, USA) was used to plot graphs by depicting the means with central values and s.e.m. with error bars.

## Data Availability

The data that support the findings of this study are available from the corresponding author upon reasonable request.

## Supporting Information

Supporting Information is available from the Wiley Online Library or from the author.

## Acknowledgements

M. S. L., H. H., Sungho K., and I. P. contributed equally to this work. J.H.L thanks to You Hwan Jo, Doyun Kim, Hyunglan Chang, and Hyuksool Kwon at SNUBH for helping the animal experiments. M.S.L. would like to thank Seyong Kwon for helpful discussion; Thao Nguyen for aiding visualization of figures; Daeho Kim for assistance in PCR experiments; Hye-Rim Sim for microbiology assistance; Soo-ah Park at the UNIST In Vivo Research center (IVRC) for supporting animal care; Jin-hoe Hur and Hong-chan Joung at the UNIST Optical Biomed-Imaging Center (UOBC) for technical advices. This work was supported by the Bio & Medical Technology Development Program of the National Research Foundation (NRF) funded by the Korean government (MSIT) (Grant No. NRF-2017M3A9E2062136, 2017M3A9E2062138, 2017M3A9E2062181, 2017M3A9E2062210). T.K. and J.H.K. thank UNIST for a research fund (1.200094.01).

## Author contributions

M.S.L. designed the µFISH system, carried out the µFISH experiments, and analyzed the data.; H. H. carried out the 16S rRNA sequencing experiments and performed data analysis, including in silico probe design validation.; I.P. designed and conducted the animal experiments and clinical study with assistance from D.-H. J. and Seonghye K.; I.P. and J.H.L analyzed the data; Sungho K. prepared and characterized the FISH probes with assistance from J.I.; H.K., J.H.L., T.K., and J.H.K. conceived the idea, led efforts to design the project, and wrote the manuscript with input from all authors.

