## Supporting Information for "Quantitative fluorescence in situ hybridization (FISH) of magnetically confined bacteria enables rapid determination of early-stage human bacteremia"

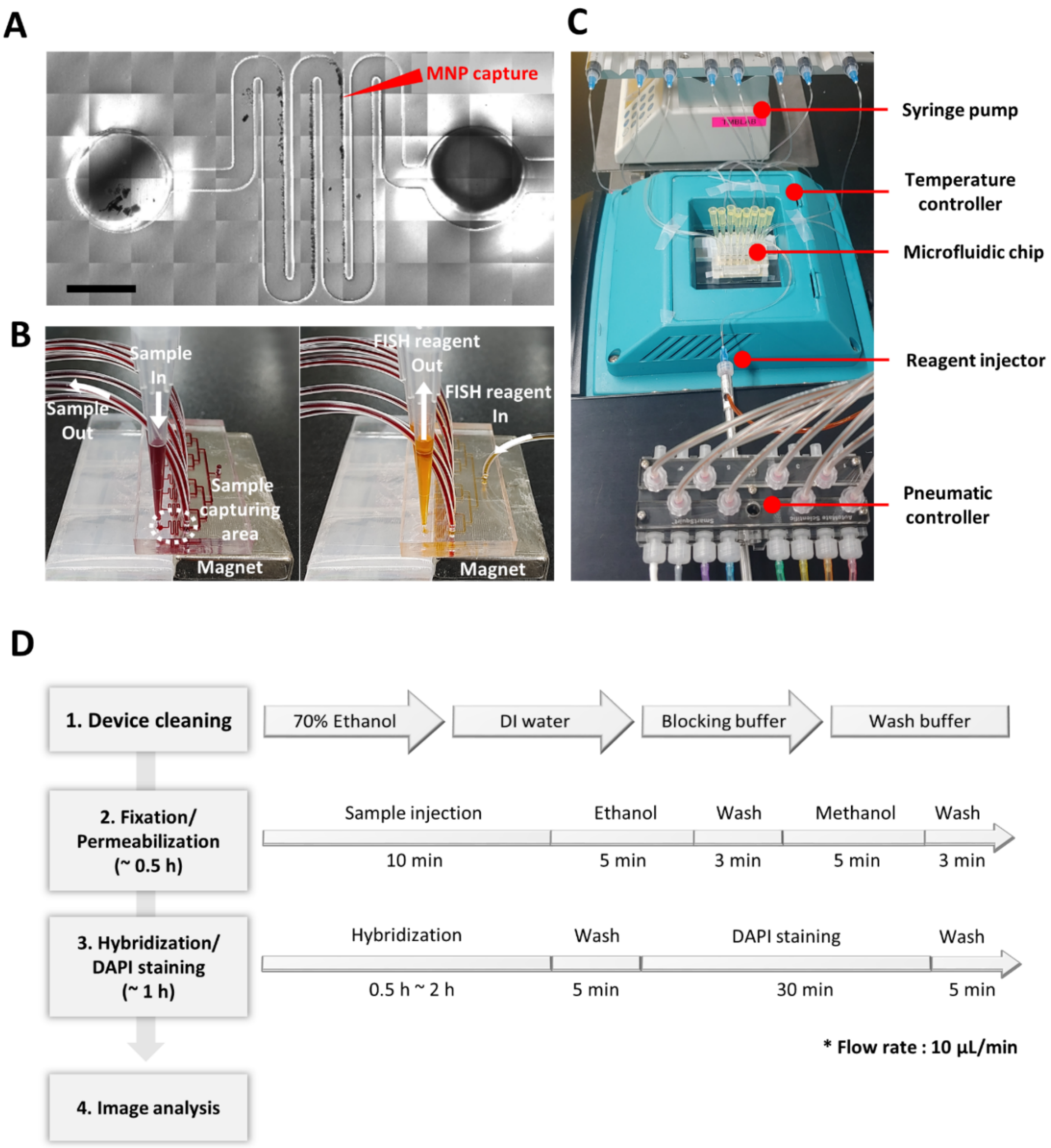


**Figure S1.** A) A stitched differential interference contrast (DIC) image showing an entire region of the μFISH device consisting of an inlet, an outlet, and a sinuous microfluidic channel. The MNPs are magnetically captured in the channel. Scale bar, 1 mm. B) The eight μFISH device units were designed in parallel to increase the throughput of μFISH analysis.

The blood samples containing MNPs were pulled from a reservoir of a pipette tip into the µFISH microfluidic channel by a syringe pump, which was connected with Tygon® tubing. The permanent magnet is located underneath the µFISH device to hold the MNP-captured bacteria on the channel surface. Reagent solutions for carrying out FISH are injected through the inlet (FISH reagent in), and waste solutions are discarded into the reservoir of the pipette tip. C) The experimental setup showing the µFISH device placed on the temperature controller, and the reagent solutions are pneumatically controlled by the automatic pressure controller. D) A flowchart describing the sequential steps for performing µFISH in the device.

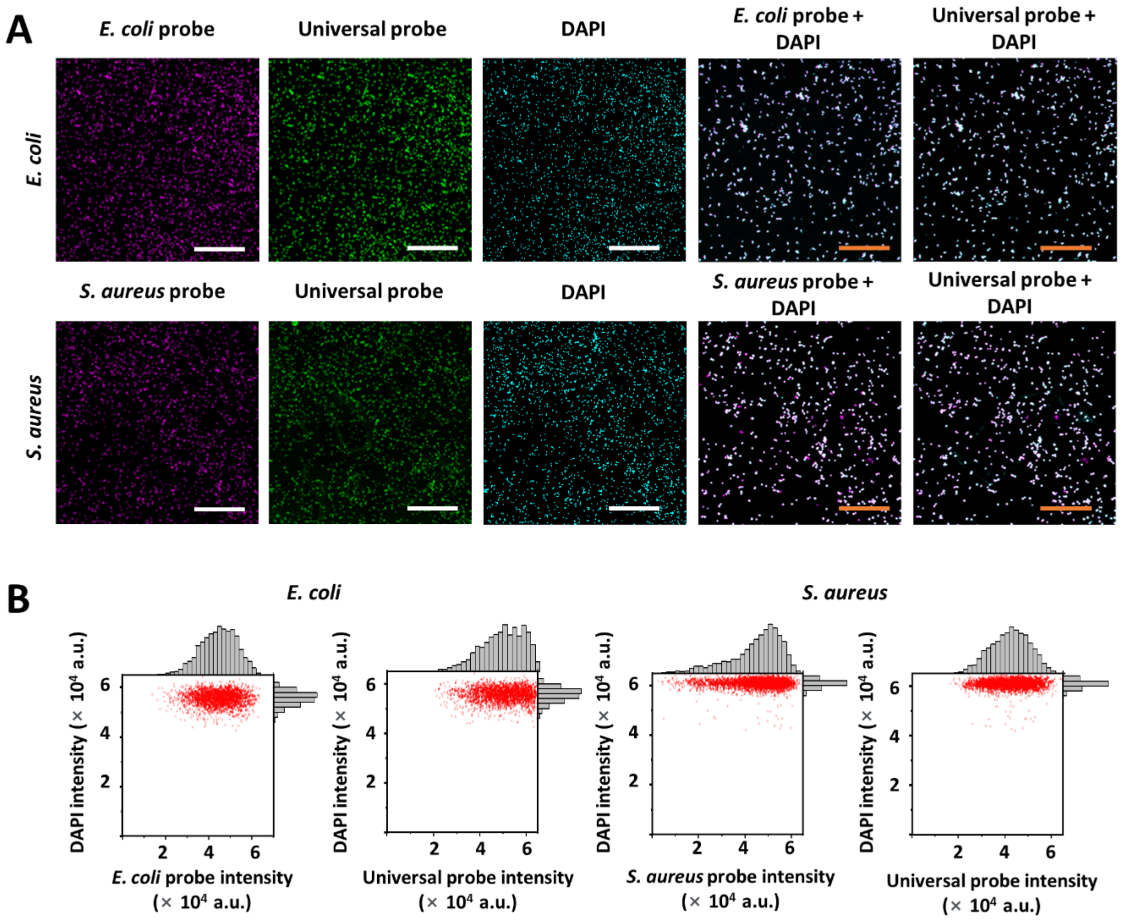


**Figure S2.** A) Fluorescence images of *E. coli* and *S. aureus* labeled with FISH probes targeting each species (magenta), universal probe (green), and DAPI (cyan). FISH and DAPI images were turned into magenta and cyan, respectively, overlaid, and shown in 2X larger magnification. Bacterial cells detected both with FISH probes and DAPI appear in white. Scale bars, 50 µm (white), 25 µm (orange). B) Scatter plots of FISH and DAPI signal intensities for the species-specific probes and the universal probe, shown for both *E. coli* (left) and *S. aureus* (right).

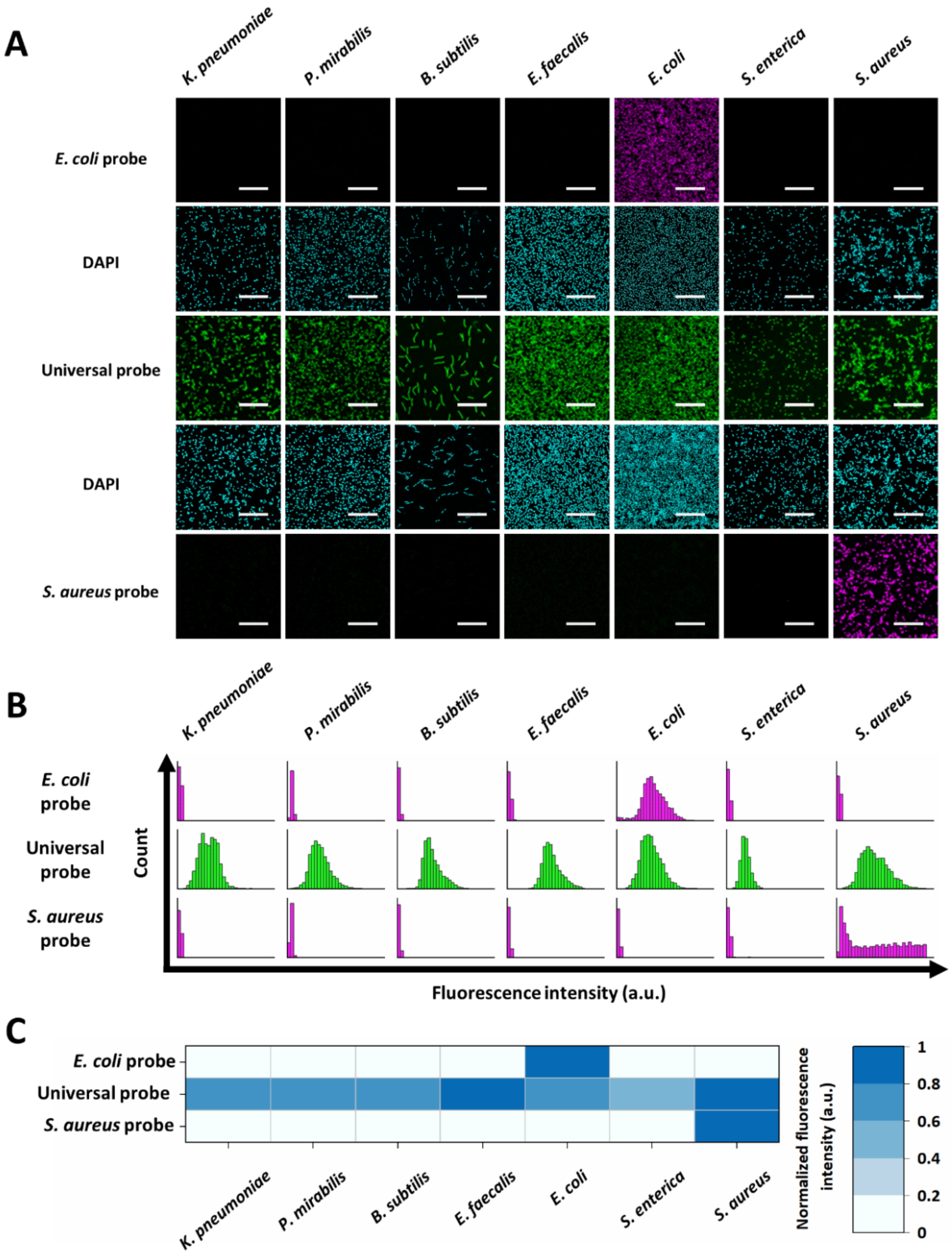


**Figure S3.** A) Fluorescence images of seven bacterial species labeled with *E. coli*-specific, *S. aureus*-specific, and universal FISH probes for the validation of target specificity. Scale bars, 20 µm. DAPI images are from the same areas as those for the specific-specific probes. B) Histograms of fluorescence signal intensity in each bacterial cell from 3,000 bacterial cells in multiple FISH images. Bacterial cells were detected based on the DAPI signal. C) A heat map showing the average fluorescence intensity from the histograms in (B). The intensity was normalized to the maximum in each row.

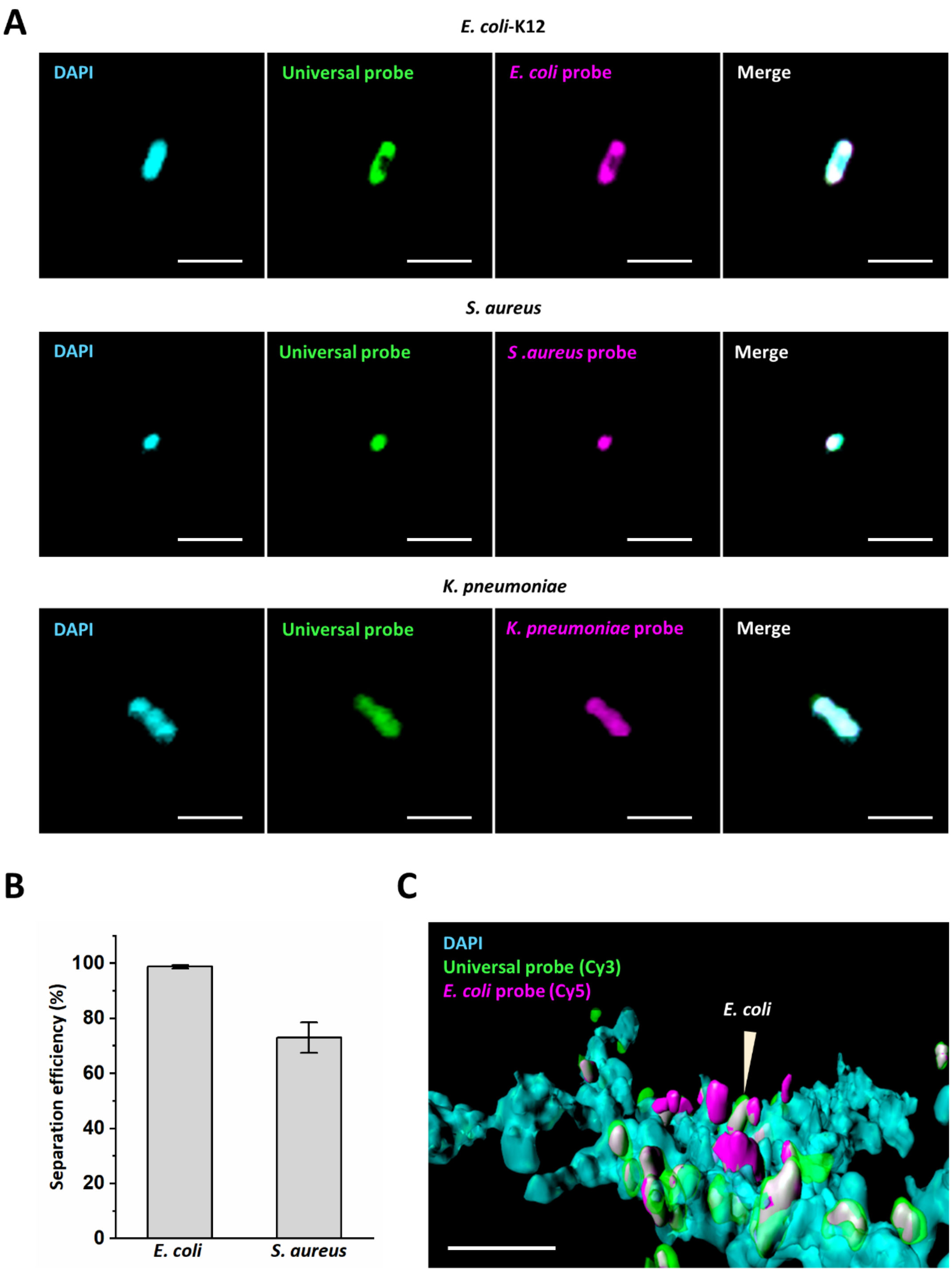


**Figure S4.** A) Magnified views of bacteria species (*E. coli*, *S. aureus*, and *K. pneumoniae*), which were labeled with DAPI, the universal probe (Cy3), and the species-specific probes (Cy5), respectively. Scale bars, 3 µm. B) The magnetic separation efficiency of *E. coli* and *S. aureus* from blood using hrMBL-MNPs ($n$ = 3). Error bars, s.e.m. C) The confocal microscopic µFISH images of bacteria-MNPs that magnetically sequestered in the µFISH device. The bacterial cells both in magenta (Cy5) and green (Cy3) represent *E. coli,* and the green bacterial cells (Cy3) are presumed bacterial species other than *E. coli*. The magnetically enriched samples also contained immune cells (DAPI) due to the phagocytosis of opsonin-coated magnetic nanoparticles. Scale bar, 10 µm.

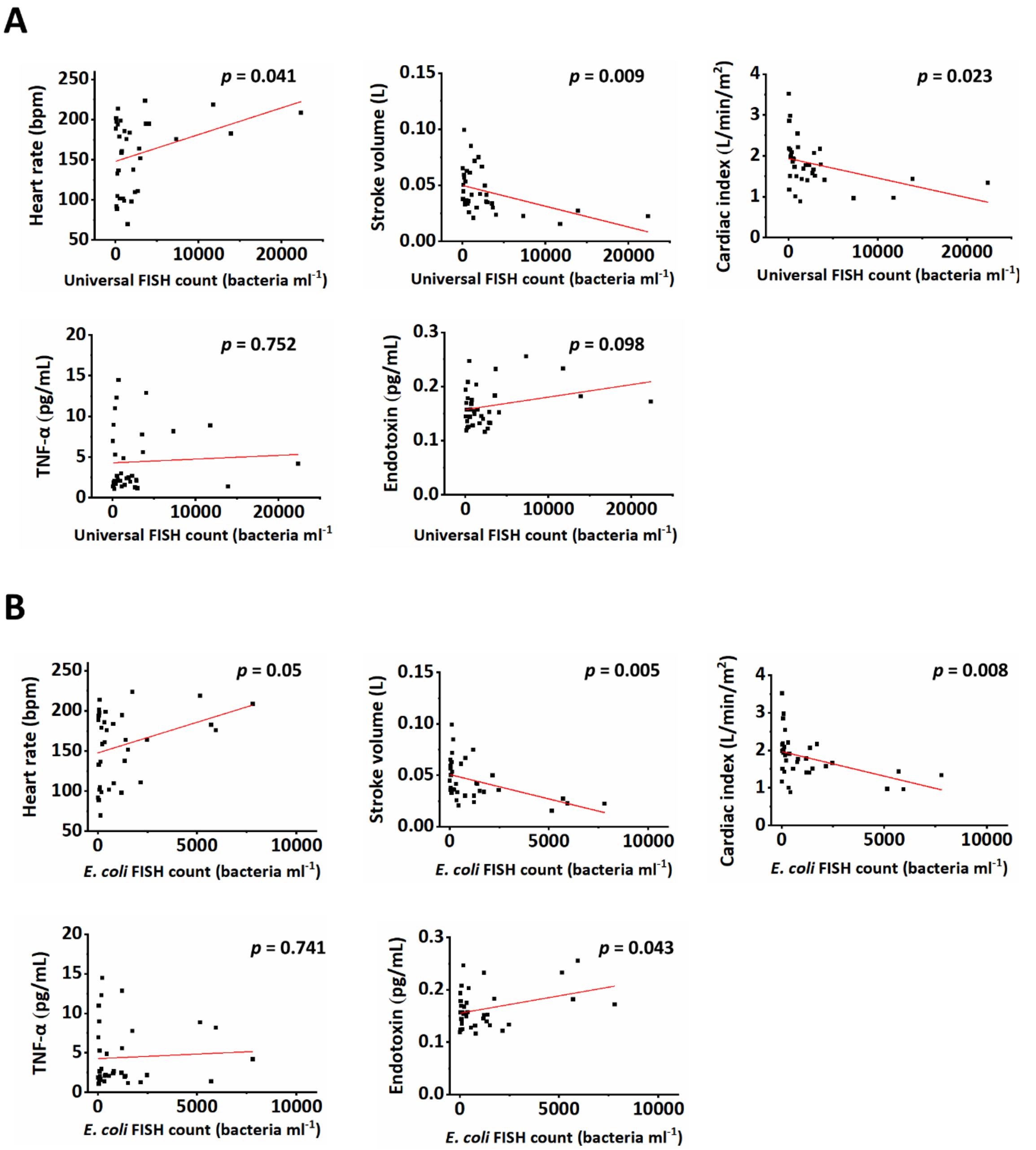


**Figure S5.** Correlation between the FISH signal counts and the observed variables in the porcine bacteremia model. A) Bacterial loads determined by the FISH signal counts of the universal probe are significantly correlated with the hemodynamic variables, such as heart rate, stroke volume, and cardiac index ($p < 0.05$, respectively), but not with TNF-α and endotoxin levels. B) The FISH signal counts of the *E. coli*-specific probe were significantly correlated with the endotoxin levels in addition to correlation with hemodynamic variables ($p < 0.05$, respectively).

**Table S1.** Demographic and clinical characteristics of the patients. UT, urinary tract.

| Variables | Patients | | | | | | | | | |
|---|---|---|---|---|---|---|---|---|---|---|
| | 1 | 2 | 3 | 4 | 5 | 6 | 7 | 8 | 9 | 10 |
| **Age range** | 66-70 | 81-85 | 76-80 | 61-65 | 76-80 | 41-45 | 76-80 | 51-55 | 46-50 | 81-85 |
| **Sex** | Female | Male | Female | Female | Female | Male | Female | Male | Female | Male |
| **Primary site of infection** | UT | Biliary tract | UT | UT | UT | UT | UT | UT | Genital tract | Biliary tract |
| **Clinical diagnosis** | Acute pyelonephritis | Acute cholangitis | Acute pyelonephritis | UT infection | Acute pyelonephritis | UT infection | Cystitis | Acute pyelonephritis | Tubo-ovarian abscess | Acute cholecystitis |
| **Initial vital signs** | | | | | | | | | | |
| Blood pressure (mmHg) | 105/37 | 142/63 | 128/66 | 131/70 | 86/46 | 130/73 | 112/64 | 170/93 | 112/69 | 129/62 |
| Heart rate (bpm) | 109 | 89 | 84 | 124 | 93 | 113 | 71 | 121 | 89 | 119 |
| Respiratory rate (cpm) | 18 | 16 | 19 | 20 | 21 | 20 | 18 | 18 | 22 | 24 |
| Body temperature (℃) | 39.9 | 39.6 | 37.8 | 38.9 | 38.2 | 38.8 | 38.5 | 39 | 38.3 | 40 |
| **Laboratory tests** | | | | | | | | | | |
| WBC (x $10^3$ $\mu L^{-1}$) | 6.8 | 7.08 | 13.92 | 0.1 | 2.2 | 12.67 | 9.22 | 10.92 | 13.43 | 12.38 |
| C-reactive protein (mg $dL^{-1}$) | 5.7 | 1.11 | 5.2 | 3.91 | 13.1 | 10.1 | 4.46 | 3.24 | 20.14 | 14.28 |
| Lactate (mmol $L^{-1}$) | 9.8 | 3.1 | - | - | 2.8 | - | 1.2 | - | - | - |
| **Microbiologic diagnosis** | | | | | | | | | | |
| Blood culture | *E. coli* | - | - | - | - | *E. coli* | - | *E. coli* | - | *K. pneumoniae* |
| Primary site culture (body fluid) | *E. coli* (urine) | *E. coli* (bile) | *E. coli* (urine) | *E. coli* (urine) | *E. coli* (urine) | *E. coli* (urine) | *E. coli* (urine) | *E. coli* (urine) | *E. coli* (pus) | *E. coli, K. pneumoniae* (bile) |
| **Clinical severity score** | | | | | | | | | | |
| SOFA score | 10 | 3 | 4 | 3 | 7 | 9 | 2 | 2 | 0 | 2 |
| APACHE II score | 26 | 9 | 15 | 9 | 20 | 8 | 9 | 9 | 2 | 11 |
